# Relationship between BrainAge Polygenetic Risk Score and plasma biomarkers in the A4/LEARN studies

**DOI:** 10.1101/2025.07.07.25331010

**Authors:** Jorge Garcia Condado, Mabel Seto, Madison Cuppels, Colin Birkenbihl, Hannah M. Klinger, Gillian T Coughlan, Michael J Properzi, Hyun-Sik Yang, Aaron P Schultz, Jasmeer Chhatwal, Dorene M Rentz, Asier Erramuzpe, Jesus M Cortes, Keith A Johnson, Reisa A Sperling, Rachel F Buckley, Ibai Diez

## Abstract

**Background and Objectives:** To examine the association between genetic predisposition to accelerated brain aging—measured with polygenic risk scores (PRS) derived from BrainAge models—and plasma biomarkers of Alzheimer’s disease (AD), with attention to age and sex-specific effects.

**Methods:** We analyzed 1994 cognitively unimpaired participants from the A4/LEARN studies (71.5±4.8 years; 41% male). We computed the genetic risk of accelerated grey matter loss associated with age using GWAS data from previous studies. Baseline plasma biomarkers included pTau_217_ (N=980; Eli Lilly immunoassay), and GFAP and NfL (N=1636; Roche Elecsys immunoassay). General linear models tested associations between each PRS and each biomarker, including PRS-by-age interaction terms. Analyses were additionally stratified by sex.

**Results:** BrainAge PRS moderated the association between age and pTau_217_ levels (β=0.08±0.03, p=0.0086), such that higher PRS for BrainAge was associated with increased pTau_217_ at older ages. Results were robust to covariates. This association was significant in females but not in males. No significant associations were found with GFAP or NfL.

**Discussion:** Genetic risk for accelerated grey matter loss with aging is associated with elevated pTau_217_ levels in cognitively unimpaired older females. These findings suggest a sex- and age-specific genetic link between brain aging and early AD pathology.

## Introduction

Individual genetic predisposition to accelerated biological aging is hypothesized to drive faster pathological accumulation and increase the risk of Alzheimer’s disease (AD). BrainAge models^1^, which estimate brain aging relative to chronological age, have been instrumental in identifying individuals exhibiting accelerated brain aging phenotypes. Furthermore, previous genome-wide association studies (GWAS) have identified polygenic combinations associated with accelerated age-related loss of grey matter, white matter, and functional connectivity^2^. This study aims to investigate the complex interplay between accelerated brain aging, genetic predisposition, and plasma biomarkers of neurodegeneration, including effects of age and sex, to elucidate the genetic underpinnings of AD-related neurodegenerative patterns.

## Methods

### Participants

Cognitively unimpaired older adults aged 65-85 enriched for elevated β-amyloid (Aβ) PET and stable medical conditions were enrolled in the A4/LEARN study. Detailed methods of the A4/LEARN study are published^3^. For this study, we selected 1994 participants with plasma biomarkers and genetic data.

### Standard Protocol Approvals and Patient Consents

We conducted the procedures for this study under the ethical guidelines stipulated by the Massachusetts General Brigham Human Research Committee, which is the relevant Institutional Review Board. Written informed consent was obtained from all participants.

### Polygenetic Risk Scores (PRS)

PRS were computed for a grey matter (GM) BrainAge model using GWAS summary statistics from Wen et al., 2024^2^. The GM model was chosen because it had the most pronounced heritability and strongest correlation with AD genetic risk^2^. PLINK 2.0 was used to calculate PRS using the *clumping + thresholding* method^4^ at a p-value threshold of 0.5. PRS were z-scored and outliers with absolute values greater than 5SD from the mean were removed. The web-based platform FUMA (v 1.5.2)^5^ was used to map GWAS summary statistics to genes and MAGMA (v1.08) for the enrichment analysis of the obtained gene set. Tissue-specific expression data was sourced from 54 tissues from GTEx v8^6^ and the GWAS Catalog^7^ was used to test for the overrepresentation of identified genes in specific phenotypes.

### Plasma Biomarkers

Plasma samples at screening were processed by two different companies: Eli Lilly and Company (Lilly) and Roche Diagnostic. Lilly conducted pTau_217_ (N=980) testing using an automated electrochemiluminescent immunoassay (Tecan Fluent workstation for preparation, MSD Sector S Imager 600MM for detection)^8^. GFAP and NfL measurements (N=1636) were conducted by Roche Diagnostics using the Elecsys Robust Prototype Immunoassays^9^. 622 participants have both analysis from Lily and Roche.

### Covariates

Age, sex and years of education were obtained during screening. *APOE*ε4 status was obtained from direct genotyping of *APOE*. Aβ status was obtained from Aβ-PET scans.

### Statistical Analysis

All statistical analyses were performed in R version 4.4.2. We first assessed group differences in PRS by sex, *APOE*ε4 status, and Aβ status using Chi-squared test, and examined Pearson’s correlations with age and years of education. In the primary analysis, we used general linear models to test associations between each of the three plasma biomarkers and the PRS, including interaction terms between age and PRS. We report standardized β coefficients. For significant models, we conducted two sensitivity analyses: (1) adjusting for age, sex, years of education, *APOE*ε4, and Aβ status, and (2) restricting to individuals with complete biomarker data and modeling all biomarkers together with full covariate adjustment. We further modeled pTau_217_ including a three-way interaction between age, PRS, and sex, and conducted sex-stratified analyses. The significance level was set at α=0.05, no multiple comparisons corrections were implemented.

### Data Availability

Data from the A4/LEARN study is available to registered researchers through Synapse^9^. Code for the analysis can be found in GitHub.

## Results

### Descriptive Results

Baseline characteristics are provided in **Table 1**. PRS scores did not differ when splitting by sex, *APOE*ε4 status or Aβ status. Further, they were not significantly correlated with age or education.

**Table 1.** Baseline Characteristics of the Study Sample (N=1994)

| Characteristic | Overall<br>N = 1,994<sup>1</sup> | Aβ-, N = 1205<sup>1</sup> | Aβ+, N = 789<sup>1</sup> | p-value<sup>2</sup> |
| --- | --- | --- | --- | --- |
| Age | 71.45 (4.79) | 71.13 (4.74) | 71.95 (4.81) | <0.001 |
| Sex |  |  |  | 0.8 |
| Female | 1,181 (59.23%) | 716 (59.42%) | 465 (58.94%) |  |
| Male | 813 (40.77%) | 489 (40.58%) | 324 (41.06%) |  |
| Education (Years) | 16.70 (2.69) | 16.73 (2.71) | 16.65 (2.66) | 0.7 |
| APOEε4 | | | | <0.001 |
| ε4- | 1,218 (61.08%) | 913 (75.77%) | 305 (38.66%) | |
| ε4+ | 776 (38.92%) | 292 (24.23%) | 484 (61.34%) | |
| Lilly Platform (N) | 980 (49.15%) | 243 (20.17%) | 737 (93.41%) | <0.001 |
| pTau217 (U/mL)<sup>3</sup> | 0.26 (0.14) | 0.17 (0.05) | 0.29 (0.15) | <0.001 |
| Roche Platform (N) | 1,636 (82.05%) | 1,185 (98.34%) | 451 (57.16%) | <0.001 |
| NfL (pg/mL)<sup>4</sup> | 3.26 (1.56) | 3.20 (1.53) | 3.41 (1.65) | 0.005 |
| GFAP (ng/mL)<sup>4</sup> | 0.10 (0.05) | 0.10 (0.05) | 0.12 (0.06) | <0.001 |

<sup>1</sup>Mean (SD); n (%)
<sup>2</sup>Wilcoxon rank sum test; Pearson's Chi-squared test
<sup>3</sup>Lilly Clinical Diagnostics Laboratory conducted pTau217 testing using an automated electrochemiluminescent immunoassay (Tecan Fluent workstation for preparation, MSD Sector S Imager 600MM for detection)
<sup>4</sup>Roche Diagnostics conducted testing of their Elecsys Robust Prototype Immunoassays.

### Primary Analysis

There was an interactive effect of PRS and baseline age on pTau_217_ levels (β=0.08±0.03, p=0.0086). That is, older age and higher PRS for BrainAge was associated with higher pTau_217_ levels. In sensitivity analyses, the PRS × age interaction remained significant after: (1) additionally adjusting for age, sex, education, *APOE*ε4, and Aβ status (β=0.13±0.04, p<0.001), and (2) including only participants with all three plasma biomarkers and adjusting for all covariates and plasma biomarkers (β=0.08±0.04, p=0.046). **Figure 1A** shows no significant correlation between PRS and pTau_217_; **Figure 1B** shows the interaction between PRS and age on pTau_217_ levels.

**Figure 1.**
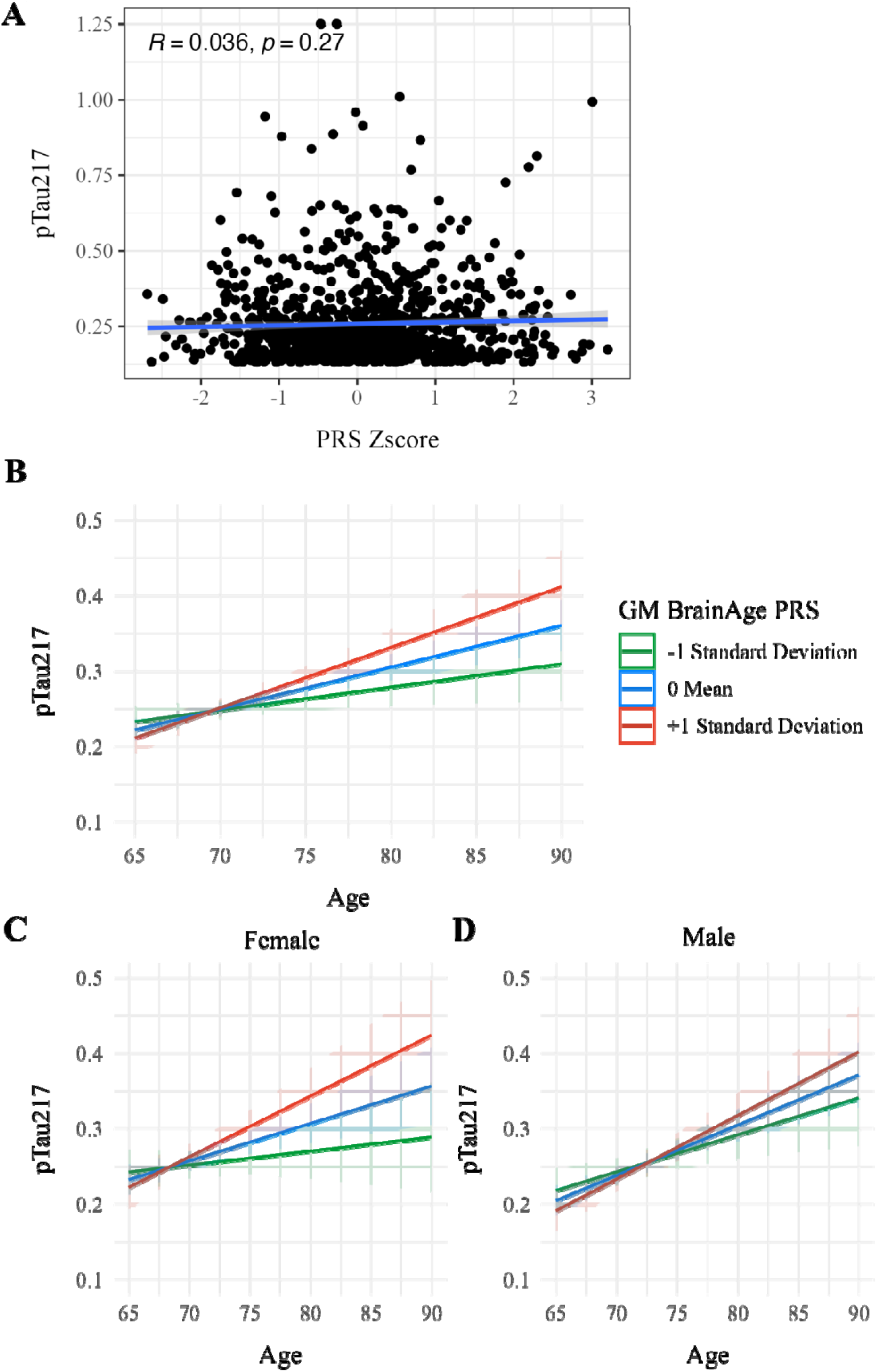
Plasma pTau_217_ levels across age for different levels of Grey Matter (GM) BrainAge Polygenic Risk Scores (PRS). **(A)** Scatter plot of PRS and pTau_217_ in cognitively normal individuals from the A4/LEARN study (N=960). **(B)** Plot of the interaction between age and PRS on pTau_217_ levels. Higher PRS (red line) is associated with a steeper increase in pTau_217_ levels with age, while lower PRS (green line) shows minimal change over time. These results indicate that PRS interacts with age to influence the trajectory of pTau_217_ levels (β = 0.085, p = 0.0086). **(C)** Stratified model in females showing a significant interaction between PRS and age (β = 0.098, p = 0.027). **(D)** Stratified model in males showing no significant interaction (β = 0.070, p = 0.14). Shaded regions represent 95% confidence intervals.

### Sex Analysis

Age was strongly associated with pTau_217_ levels in males (β=0.27±0.05, p<1×10□□), with a weaker effect observed in females (β=0.16±0.04, p<0.001). Given these sex-based differences we investigated potential interactions. The three-way interaction between PRS, age, and sex on pTau_217_ was not significant. Consequently, we explored sex-stratified models. In females, the PRS × age interaction was significant (β=0.10±0.04, p=0.027), while in males, the direction was consistent with that seen in females but weaker and not significant (β=0.07±0.05, p=0.14). **Figure 1C** and **1D** show the stratified models for males and females, respectively.

### Genetic Enrichment Analysis

The gene set enrichment analysis in PRS revealed significant overlap with several neuropsychiatric and neuroanatomical traits as seen in **Figure 2**. Among the identified genes, one of the strongest associations was observed for *MAPT*, encoding the tau protein, which is heavily implicated in AD and other neurodegenerative disorders^10^. Tissue expression analysis showed that the associated genes were predominantly expressed in brain regions, consistent with the PRS targeting grey matter.

**Figure 2.**
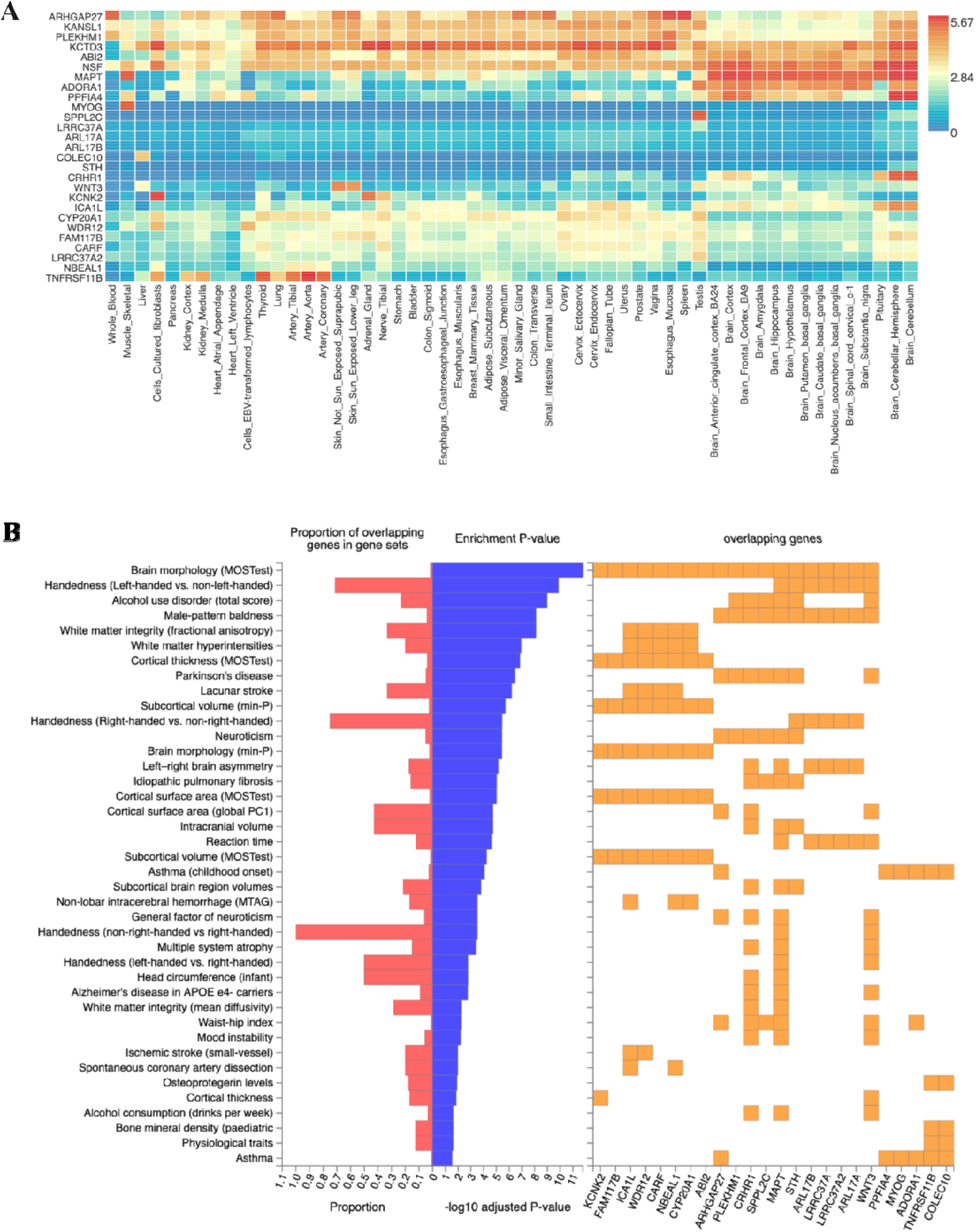
Genetic Drivers of Grey Matter BrainAge Polygenetic Risk Scores (PRS) **(A)** Heatmap depicting GTEx 54 tissue-specific expression of genes associated with the PRS. Color intensity indicates normalized average gene expression levels across various human tissues (columns), highlighting tissues with enriched expression of PRS-related genes. **(B)** Gene set enrichment analysis revealing biological pathways and complex traits genetically linked to the PRS. The left panel shows the proportion of overlapping genes with trait-associated sets, while the middle panel displays the significance of these overlaps (-log10 adjusted p-value). The right panel illustrates specific genes overlapping with GWAS hits for other traits, indicating potential pleiotropy.

## Discussion

In this study, a higher genetic predisposition to accelerated grey matter loss captured by BrainAge PRS was associated with increased pTau_217_ levels with age. Plasma pTau_217_ is known to be more strongly associated with grey matter neurodegeneration, which further supports the specificity of this finding^11^. The link between PRS and age-related biomarker changes aligns with broader research showing associations between PRS and age gaps in other organ systems^12^, indicating a promising avenue for understanding systemic aging-related vulnerability.

In sex-stratified analyses, a significant BrainAge PRS × age interaction on pTau_217_ levels was observed in women but not in men. In women, the interaction suggests that genetic risk may modulate age-related increases in tau pathology. However, since this analysis does not test for interaction effects, we cannot conclude that the effect differs by sex; the same pattern could exist in men but remain undetected due to limited power. These findings align with prior reports that pTau_217_ more strongly predicts cognitive decline and brain atrophy in cognitively unimpaired women^13^, supporting the possibility that women may be more susceptible to AD-related tau pathology.

The genetic enrichment analysis identified *MAPT* as one of the strongest associations within the BrainAge PRS-linked gene set, consistent with previous studies^14^. Given that *MAPT* is involved in tau production^10^, its prominence in the PRS suggests a potential genetic link between grey matter BrainAge models and pTau_217_ levels. This points to *MAPT* genetic variation as a potential contributor to tau pathology in individuals with higher BrainaAge PRS, possibly through altered isoform expression^15^.

In conclusion, our findings show that BrainAge PRS is associated with pTau_217_ levels in an age- and sex-dependent manner. This highlights the relevance of genetic background and demographic factors in neurodegenerative risk. A key limitation of this study is the use of a single dataset with limited sample size and low racial diversity, particularly in stratified analyses, which may reduce generalizability and statistical power. Another limitation is that the PRS models explain only a small proportion of variance in BrainAge, with an R^2^ values of 0.81%^2^. A further limitation is that sex-stratified analysis cannot be interpreted as definitive sex differences, as similar effects may exist in both groups but remain undetected due to limited power. Our results support the use of polygenic scores in identifying individuals at elevated risk for AD-related pathology.

## Data Availability

Data from the A4/LEARN study is available to registered researchers through Synapse. Code for the analysis can be found in GitHub.

https://www.synapse.org/Synapse:syn61250768/wiki/628717

https://github.com/JGarciaCondado/PRSBrainAgePlasma

## Acknowledgments

The research that led to this publication was conducted with the support of a US-Spain Fulbright grant. The project that gave rise to these results received the support of a fellowship from “la Caixa” Foundation (ID 100010434) to J.G.C. The fellowship code is LCF/BQ/DI21/11860030. J.M.C acknowledges financial support from the Health Department of the Basque Country (grants 202211103 and 2023111002). M.S. is supported by a research fellowship from the Alzheimer’s Association (24AARF-1201281). G.T.C. is funded on the National Institute on Aging (K99 AG083063), as well as a research fellowship from the Alzheimer’s Association (AARF-23-1151259). A.E. is supported by the Spanish Ministry of Science and Innovation, grant RYC2021-032390-I and the Basque Country’s Department of Economic Development and Infrastructure (Elkartek Program) under Grant KK-2024/00028. J.M.C., A.E. and I.D. are supported by Ikerbasque: The Basque Foundation for Science. J.M.C and A.E. are supported by the Spanish Ministry of Science under Grant PID2023-148012OB-I00. J.M.C. acknowledges financial support from the Basque Ministry of Health (grants 2023111002 & 2022111031). R.F.B. is supported by the National Institute on Aging (R01AG079142 and DP2 AG082342). I.D. was supported by Alzheimer’s Association (AARF-23-1145358) and Spanish Ministry of Science (RYC2022-035429-I and PID2023-150633OA-I00).

The A4 study is funded by a public-private-philanthropic partnership, including funding from the National Institutes of Health-National Institute on Aging (U19AG010483; R01AG063689), Eli Lilly and Company, Alzheimer’s Association, Accelerating Medicines Partnership, GHR Foundation, an anonymous foundation and additional private donors, with in-kind support from Avid, Cogstate, Albert Einstein College of Medicine, US Against Alzheimer’s disease, and Foundation for Neurologic Diseases. The companion observational Longitudinal Evaluation of Amyloid Risk and Neurodegeneration (LEARN) study is funded by the Alzheimer’s Association and GHR Foundation.

